# Using genotyping and whole-exome sequencing data to improve genetic risk prediction in deep venous thrombosis

**DOI:** 10.1101/2022.04.24.22274229

**Authors:** Valeria Lo Faro, Therese Johansson, Julia Höglund, Fatemeh Hadizadeh, Åsa Johansson

**Affiliations:** Department of Immunology, Genetics and Pathology, Science for Life Laboratory, Uppsala University, Uppsala, Sweden; Centre for Women’s Mental Health during the Reproductive Lifespan – Womher, Uppsala University, Sweden

## Abstract

**Background:** Deep Vein Thrombosis (DVT) is a common disease that can lead to serious complications such as pulmonary embolism and in-hospital mortality. More than 60% of DVT risk is influenced by genetic factors, such as Factor V Leiden (FVL) and prothrombin G20210A mutations (PTM). Characterising the genetic contribution and stratifying participants based on their genetic makeup can favourably impact risk prediction. Therefore, we aimed to develop and evaluate a genetic-based prediction model for DVT based on polygenic risk score (PRS) in the UK Biobank cohort.

**Methods:** We performed a genome-wide association study (GWAS) and constructed a PRS in the 60% (N=284,591) of the UK Biobank cohort. The remaining 40% (N=147,164) was employed to evaluate the PRS and to perform gene-based tests on exome-sequencing data to identify effects by rare variants.

**Results:** In the GWAS, we discovered and replicated a novel variant (rs11604583) near *TRIM51* gene and in the exome-sequencing data, and we identified a novel rare variant (rs187725533) located near *CREB3L1*, associated with 2.2-fold higher risk of DVT. In our PRS model, the top decile is associated with 3.4-fold increased risk of DVT, an effect that is 2.3-fold, when excluding FVL carriers. In the top PRS decile, cumulative risk of DVT at age of 80 years is 10% for FVL carriers, contraposed to 5% for FVL non-carriers.

**Conclusion:** We showed that common and rare variants influence DVT risk and that the PRS improves risk prediction on top of FVL. This suggests that individuals classified with high PRS scores could benefit from early genetic screening.

## INTRODUCTION

In the last century life expectancy has doubled thanks to a global shift of morbidity and mortality from infectious to non-infectious causes, making cardiovascular disease the leading cause of death.^1^ Myocardial infarction and stroke are well recognised cardiovascular causes of death, however, among cardiovascular morbidity, venous thromboembolism (VTE) also contributes massively to the global burden of non-infectious disease.^2^ In fact, the incidence of VTE has been estimated to be approximately 10 million worldwide, annually, resulting in more than half a million deaths in Europe, which is increased by aging.^2^ In the United Kingdom, VTE alone is responsible for at least 25,000 deaths annually, and is the most common cause of hospital mortality.^3^ VTE is a disease that is caused by the formation of blood clots in the veins.^4^ When the blood clots formation occurs in a deep vein, this condition is called deep venous thrombosis (DVT) that accounts for two-thirds of all VTE cases. When the blood clots in the deep veins migrate to the lungs blocking the blood flow, this becomes a case of pulmonary embolism (PE), a condition that can be lethal. In approximately 90% of PE cases, the emboli are caused by formation of thrombosis in proximal leg deep vein.^5,6^ Common symptoms of DVT are not specific and include pain, swelling, tenderness of the affected limb and warmth at the site of the clots. However, it is often asymptomatic and if left untreated, DVT can lead to serious health problems, such as heart failure other than PE.^7^ The identification of risk factors associated with DVT is likely to lead to a reduction in the incidence, morbidity and mortality caused by this condition, especially where the risk factors are modifiable. Furthermore, the identification of causal risk factors for DVT can aim in the development of efficient prophylactic drugs.^8^

DVT is a complex condition with both acquired and genetic risk factors. Among the acquired risk factors, there are advancing age, prolonged hospitalisation, surgery, and immobilisation as well as administration of oestrogen therapy, cancer, chemotherapy, obesity, pregnancy, and smoking.^6^ Regarding the genetic factors that play a role in DVT, they are mostly mutations in the inherited thrombophilia genes, such as the coagulation Factor V and Factor II or prothrombin. Known mutations in these genes that cause thrombosis are the Factor V Leiden (FVL) mutation and the prothrombin G20210A mutation (PTM).^9^ It has been shown that prevalence of FVL and PTM is the highest in Caucasians with frequencies up to 7% and 2%, respectively, in the general European population.^10^

Twin and family-based studies estimated the heritability of DVT to be approximately 60%.^11^ Heritability was also estimated for the coagulation factors involved in clot formation, such as the coagulation Factor II (49-57%) and Factor V (44-62%) genes.^12^ Identifying mutations or rare variants can change clinical care, by tailoring diagnostic and treatment strategies.^13^ For example, treatment can be improved in participants carrying rare pathogenic variants since they may respond more appropriately to specific therapies, as shown in the case of monogenic forms of type 2 diabetes, where oral pills can give to patients more benefit than insulin therapy.^14^ Despite these advantages, rare variants are not routinely sought in the course of clinical care for most of the diseases, often because targeted sequencing panels or exome sequencing are expensive. Indeed, these are costly technologies that generally have a low impact due to the paucity of deleterious mutations in the general population. Therefore, development of new approaches to catch carriers of mutations causing disease would improve clinical care, for example by identifying high-risk individuals suitable for further sequencing analysis.

One way to identify individuals with high genetic liability to develop DVT is through polygenic risk scores (PRSs). PRSs are calculated from common genomic variants that an individual carries across the whole genome, weighted by the effect sizes that have been obtained by regression analysis. Rare genetic variants are not commonly captured by the PRS since are not in linkage disequilibrium with common variants.^15^ However, the PRSs have the potential to identify a substantially larger fraction of individuals at greater disease risk in a population compared to the research of rare monogenic mutations by clinical routine. Whereas identifying carriers of rare monogenic mutations requires sequencing of specific genes and careful interpretation of the functional effects of mutations found, PRSs can be readily calculated using data generated by a single genotyping array. Benefits of identifying individuals at high risk of developing a disease can give multiple prevention lines. First, avoiding the disease with primary prevention can be accomplished by a change in lifestyle or encouraging to start proactive therapy. Second, detecting a disease at an early stage and preventing it from worsening with secondary prevention could be accomplished through more frequent screening of those at high risk. Third, a better understanding of treatment response, particularly responses to prescribed drugs, could help with tertiary prevention, which aims to improve quality of life or reduce symptoms.^16^

As said, the PRS assesses common genetic variations in millions of single nucleotide polymorphisms (SNPs), with reduced costs.^17^ Large cohort resources that conducted genome-wide association studies (GWAS) have identified genetic risk factors in loci of genes involved in the coagulation cascade, like *F2, F5, F11, FGG, VWF*, and *ABO*.^18,19^ These findings can be translated into clinical utility to estimate the individual genetic predisposition to the disease.^20– 22^ In a study conducted on individuals affected by cardiovascular diseases from UK Biobank, Sun and co-authors quantified the clinical utility of PRSs on top of conventional risk factors. They reported that the addition of PRS leads to the prevention of 7% more events than only conventional risk factors, suggesting an efficient use of the PRS.^23^ Another study showed that PRS improves the prediction of myocardial infarction in early in life, when other risk variables had not yet manifested or unavailable.^24^ Given promising effects, many large health-care systems have initiated research programs by genome-wide genotyping large proportion of population.^25,26^

Currently, most genetic investigations have focused on genetic risk factors for VTE, which include both DVT and PE cases.^27–29^ Hence, there is still a necessity to investigate the genetics underlying DVT alone and its risk prediction in the population, since this can help to prevent consequences of DVT, such as the formation of emboli migrating in the circulatory system. Indeed, the evaluation of the genetic risk factors can be used to prevent DVT in high-risk patients, e.g., through the administration of thromboprophylaxis treatment in specific situations where participants could be more susceptible or exposed to higher risk.

In this study, using both genotyping and whole-exome sequencing (WES) data from the UK Biobank (UKB), our aims were to (i) identify genetic associations for DVT using both GWAS and gene-based tests for associations, (ii) construct PRS for DVT, and (iii) evaluate the performance of the PRS to improve risk prediction in DVT.

## METHODS

### UK Biobank

We used the UK Biobank (UKB), one of the largest health cohort studies to date. During 2006– 2010, the UKB recruited more than 500,000 participants, aged between 40 and 70 years, from multiple assessment centres located in the United Kingdom, and collected a wide range of phenotype information and biological samples, such as blood samples for DNA extraction. In the UKB, phenotype information has been collected through questionnaires, interviews, and hospital records during participants recruitment visits. In addition, UK Biobank is integrating each participant’s electronic health records into their database, including inpatient International Classification of Diseases (ICD-9 and 10) diagnosis codes. For genetic analysis, the UKB conducted and released high-quality genome-wide genotyping to the researchers who have had an application for its access. The UKB analysis performed in this study has been approved by the research ethics committee (reference 11/NW/0382) and the analysis performed in this study was approved by the UKB (application #41143) and the Swedish Ethical Review Authority (Dnr: 2020-04415).

### Deep vein thrombosis data

To assess information regarding DVT disease status for the UKB participants, information was taken from different categories: main and secondary diagnoses made during hospital stay, medical conditions assessed from both verbal interviews and touchscreen questionnaires. Here, DVT was ascertained at baseline by self-report, followed by a verbal interview with a trained nurse at one of the Biobank Assessment Centres, in order to confirm diagnosis (20002:1094) and hospitalisation report following the ICD-9 451 and ICD-10 Code 180.1,180.2, and I82.2. For DVT controls, excluded criteria for their selection were those without ICD-9 Code 451, and ICD-10 Codes I80.1, I80.2, I82.2, D68, O87, O223, and including all those passing quality control in the genetic analysis.

### Genotyping quality controls and genome-wide association study

A total of 438,417 UKB participants had been genotyped using the UKB Axiom array, and another 49,994 participants had been genotyped using the similar UK BiLEVE array. The third release of imputed genotypes from the UK Biobank, data that had already been imputed using a combined 1000 Genomes/UK10K reference panel, were used for the current study. Variant level quality controls exclusion criteria included call rate < 95%, Hardy-Weinberg equilibrium *p* <1× 10^−6^, SNPs with a minor allele frequency ≤ 1%, and imputation quality ≤ 0.3. We excluded participants who were not classified as Caucasian by principal component analysis. This left a total of 482,952 participants for our downstream analyses.

One part of the cohort, including 60% of the participants (N=284,591), was used for GWAS and for constructing the PRS (training cohort), and remaining participants were used for testing the performance of the PRS (target cohort). In the GWAS, we used a binary DVT variable as outcome where cases were classified as participants that had received a DVT diagnosed prior to the end of the follow up (February 2021). Following covariates were included: age when attended assessment centre (UKB Fields 21003), sex (UKB Fields 31), the first 10 genetic principal components, and genotyping array. The SNPs were modelled in an additive model (carriers of 0, 1, or 2 copies of effect allele). We utilised SAIGE (version 0.44.5) software to perform logistic mixed models, given its ability to adjust for population structure and relatedness in population-based cohorts, and the genetic relationship matrix was constructed using 645,544 genotyped variants.^30^ The canonical Bonferroni correction for genome-wide significance, corresponding to *p* < 5 × 10^−8^ was applied. To characterise risk loci of GWAS, we clumped independent significant SNPs and defined genomic risk loci using the software PLINK v1.90. ^31^

To replicate DVT’s significant novel GWAS hits, we used the UKB target cohort (the target cohort) and FinnGen, an independent cohort of individuals of Finnish ancestry. Briefly, FinnGen is a large public-private partnership that aims to identify genotype–phenotype correlations in approximately 500,000 Finnish participants. Patients and controls in FinnGen provided informed consent for biobank research, based on the Finnish Biobank Act (https://www.finngen.fi/fi). The FinnGen, for the disease category “IX Diseases of the circulatory system (I9_)” and phenotype “DVT of lower extremities” included 5,632 cases and 225,735 controls.

### Whole exome sequencing and gene-based tests to identify rare variant associations

We used the UKB200K release of the UKB WES data, in which there are a total of 198,362 participants.^25,32,33^ Exome sequencing had been performed using IDT xGen Exome Research Panel v1.0 on the Illumina NovaSeq 6000 platform, as described previously.^25,32,33^ In order to reduce the influence of population stratification, which could be due to some rare variants being a population or even family-specific, we only included participants who self-reported being of white British descent, and who were classified as Caucasians by principal component analysis. We also excluded first-, and second-degree relatives using genetic kinship (IBS < 0.044), resulting in 147,164 participants with WES data included in the gene-based analysis. To identify rare variants associated with DVT status, we performed the gene-based SKAT-O tests using SAIGE-GENE.^30^ For the SKAT-O analysis, variants were weighted by their minor allele frequency (MAF), according to the default β (1,25) density function, as described elsewhere.^34^

The WES variants were annotated using ANNOVAR version 2017.07.16. For the SKAT-O analyses, we included all protein-altering variants, therefore excluded intronic, synonymous and non-frameshift variations, and tested a total of 22,781 genes, with at least one protein-altering variant after filtering. Bonferroni correction for the number of genes and tests performed was used to set the significance threshold for the gene-based analysis corresponding to *p* < 8.7 × 10^−7^ (0.05/22,781 tests). Conditional analysis was performed to condition on the most significant SNP (from our GWAS), within 500 kB of the gene. The same covariates as in the GWAS were also used in the SKAT-O analyses.

### Construction of Polygenic risk score and estimate of hazard ratios

PRSs were generated using polygenic risk scores–continuous shrinkage (PRS-CS) with the summary statistic of our GWAS (performed in 60% of the UKB cohort) as input.^35^ PRS-CS uses a high-dimensional Bayesian framework to model linkage disequilibrium and a continuous shrinkage prior on SNP effect sizes. We used the 1000 Genomes Project Phase 3 European sample as the LD reference.^36^ Only SNPs with a MAF ≥ 1% and imputation quality ≥0.3 were used for construction of the PRS, resulting in 8,739,041 SNPs.

A non-overlapping set of 40% of the UKB participants (target cohort) were selected to test the performance of the PRS. We excluded first- and second-degree relatives using kinship data, by using a cut-off for the estimated kinship (0.044). All individuals in the test set were scored, and the PRSs were standardised to have mean=0 and standard deviation =1. The area under curve (AUC) was calculated to evaluate the discriminative ability of PRS model. Receiver operating characteristic curve (ROC) was plotted to evaluate sensitivity and specificity of different analysed models. Finally, the PRSs were categorised into decile groups with the first decile (the 10% of the test set with the lowest PRS) considered as the reference category in all statistical tests. We evaluated the performance of the PRS in identifying individuals of high risk of developing DVT.

In our analysis, we also included a Cox regression. Here, in the Cox regression model, we used unrelated participants and the DVT hazard ratio (HR) was estimated with the standardised quantitative PRS as exposure. For DVT cases, the date when the DVT event was first reported was provided by the UKB. We utilised age as the time scale representing the time-to-event. For cases we took into account the age at diagnosis, for the controls the age at last assessment. Age at diagnosis was estimated by calculating the difference between when disease event was first reported (Data-Field 131396, for ICD 10: I80) and year of birth (Data-Field 34), rounded off downward. This left out the remaining 3,735 cases and 141,317 controls. We estimated the hazard ratios (HR) and its 95% confidence interval (CI) associated with DVT risk. Violation of the proportional hazard assumption, such as relative hazard not constant over time with different covariates, was assessed by inspecting the Schoenfeld residuals using the function cox.zph() in R.^37^ In the main analysis, the PRS was treated as a continuous covariate, and we estimated the HR per unit of the PRS. For our purpose, we used “survminer” package in R.^38^

## RESULTS

### Study design

In this study, we focused on genetic risk prediction in UKB participants of white British ancestry for which genotyping and exome sequencing data were available. We divided the white British cohort into two data sets: (1) a training set including 284,591 participants (a total 8,231 DVT cases and 276,360 DVT free controls) to construct the PRS; (2) a target set including 147,164 participants (a total 4,342 DVT cases and 142,822 DVT free controls) for testing the PRS. The target set, from which the exome sequencing data was available for all the participants, was also used to identify rare pathogenic variants. Characteristics of these two data sets are provided in Table 1.

**Table 1.** Characteristics of the training and target cohort in the UKB cohort, reported as median, interquartile range (IQR), and percentage.

|  | Training cohort |  | Target cohort |  |
| --- | --- | --- | --- | --- |
|  | *DVT cases*<br>(*n*=8,231) | *DVT controls*<br>(*n*=276,360) | *DVT cases*<br>(*n*=4,342) | *DVT controls*<br>(*n*=142,822) |
| Age (median [IQR]) | 61 [55, 66] | 58 [50, 63] | 61 [56, 65] | 58 [51, 63] |
| Female, *n* (%) | 4,460 (54.1) | 148,075 (53.5) | 2,364 (54.4) | 78,036 (54.6) |

### Association analysis

We performed a GWAS on the participants included in the training cohort. Here, the single variant association analysis showed no sign of genomic inflation (lambda= 1.05). A total of 11 genome-wide significant loci were identified (Figure 1, Table 2). Among these, two novel loci were identified, near the genes *TRIM51* (rs11604583, effect allele G, odds ratio [OR]: 0.84; 95% confidence interval [CI]: 0.79-0.89; *p* =9.81× 10^−9^) and *SLX41P* (rs73078758, OR:1.29; 95%CI: 1.18- 1.42; *p*=4.2× 10^−8^). Of these two loci, the former was replicated in the UKB target cohort (OR:0.87; 95%CI 0.8-0.95; *p* =0.001) and in the FinnGen (rs11604583, allele reference G, OR:0.88; *p*=4.0× 10^−3^). Furthermore, in our analysis the strongest association was observed for rs6025 at the *F5* locus (effective allele C, OR: 0.20; 95%CI: 0.17–0.22; *p* = 4.62 × 10^−140^), which is also the SNP for which the T-allele is the well-known FVL. In addition, we observed significant associations for other previously known genes, *ABO, F2, F11, FGA* and *VWF* genes (Figure 1, Table 2), for which the lead-SNP at *F2* was the well-known PTM.

**Figure 1:**
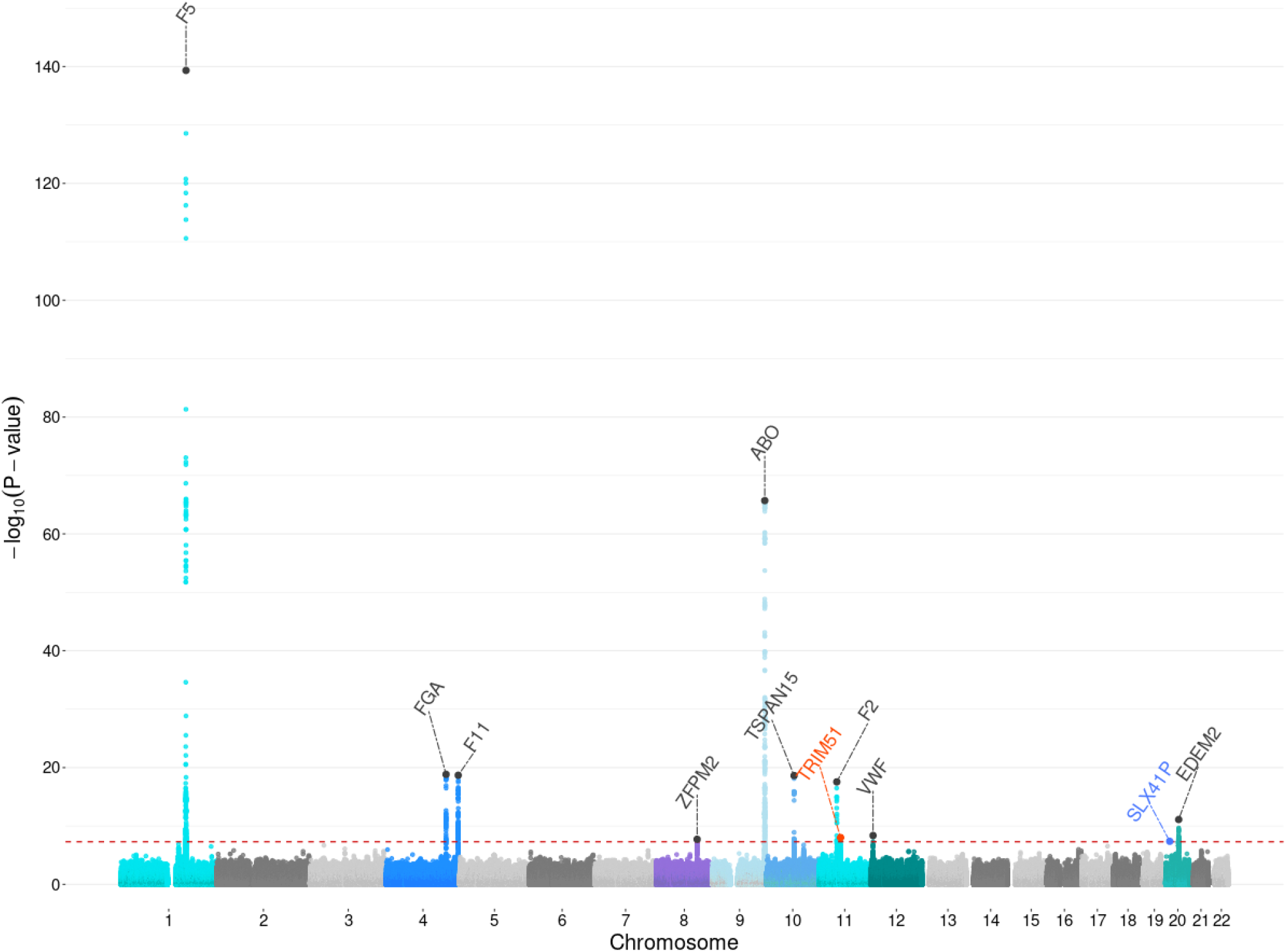
Manhattan plot for the UKB DVT analysis in the training cohort. Each dot represents a SNP, the x-axis shows the chromosomes where each SNP is located, and the y-axis shows -log10 P-value of the association of each SNP with DVT. The red horizontal line shows the genome-wide significant threshold (P-value = 5e-8; -log10 P-value = 7.30). The 11 regions that reach the threshold for significance are indicated by an annotation of the most likely causal gene in each region. The novel locus identified and replicated is highlighted in orange. The locus that could not be replicated is highlighted in blue.

**Table 2:** All the significantly associated loci associated with DVT identified in the training cohort. Chromosome locations, allele frequencies, effect sizes, and p-values of the lead variant are shown. In bold are highlighted the novel loci identified in this study.

| Leading variant | Position | P-value | Gene closest | Allele1 | Allele2 | Allele2 frequency | Odds ratio | 95% Confidence interval |
| --- | --- | --- | --- | --- | --- | --- | --- | --- |
| rs6025 | 1:169519049 | 4.62E-140 | *F5* | T | C | 0.98 | 0.20 | 0.17-0.22 |
| rs13109457 | 4:155514879 | 1.44E-19 | *FGA* | G | A | 0.25 | 1.18 | 1.13-1.22 |
| rs4253417 | 4:187199005 | 2.06E-19 | *F11* | T | C | 0.40 | 1.16 | 1.12-1.19 |
| rs16873346 | 8:106549481 | 1.90E-08 | *ZFPM2* | G | C | 0.23 | 0.90 | 0.8-0.93 |
| rs687289 | 9:136137106 | 1.99E-66 | *ABO* | G | A | 0.32 | 1.35 | 1.3-1.39 |
| rs78707713 | 10:71245276 | 2.37E-19 | *TSPAN15* | T | C | 0.12 | 0.80 | 0.76-0.84 |
| rs1799963 | 11:46761055 | 2.85E-18 | *F2* | G | A | 0.01 | 2.06 | 1.75-2.42 |
| **rs11604583** | **11:56596337** | **9.81E-09** | ***TRIM51*** | **C** | **G** | **0.08** | **0.85** | **0.79-0.89** |
| rs7308002 | 12:6152727 | 4.29E-09 | *VWF* | G | A | 0.37 | 1.10 | 1.06-1.13 |
| **rs73078758** | **20:10523896** | **4.21E-08** | ***SLX41P*** | **A** | **G** | **0.03** | **1.30** | **1.18-1.42** |
| rs1217695516 | 20:33724557 | 7.85E-12 | *EDEM2* | GAA | G | 0.14 | 1.18 | 1.12-1.23 |

In the gene-based SKAT-O analysis with the WES data, three genes *F5, F2* and *CREB3L1* genes reached the significance level (*p* of 2.83× 10^−79^, 1.23× 10^−22^, and 1.04 × 10^−17^, respectively). The most strongly associated SNPs in these genes were rs6025 in *F5* [OR (T-allele): 2.7; 95%CI: 2.5– 3.08; *p* = 1.58 × 10^−78^] and rs1799963 for *F2* (OR: 2.16; 95%CI: 1.85–2.52; *p* = 8.9 × 10^−23^). These two variants are the FVL and PTM; in our dataset 6,872 participants were carrying any of these two pathogenic variants, of which 668 have had a DVT diagnosis. The most strongly associated rare variant (MAF threshold < 0.01) observed was in the *CREB3L1* gene (rs187725533, OR:2.47; 95%CI: 2–3.06; *p* = 7.51 × 10^−17^), a missense variant with MAF = 0.005, and identified in 112 DVT cases.

### Polygenic risk score associated with deep venous thrombosis risk

For the UKB participants that were not included in the training cohort GWAS (40% of the total cohort), the PRS performance was evaluated. In the training cohort, the AUC estimated, for a model that contained the PRS in addition to the covariates of the base model (age and sex), was 0.64 (95% CI: 0.63–0.65), (Figure 2A). Furthermore, the AUC was calculated to estimate how the risk model with the PRS and base model discriminated against only the base model. The AUC of PRS and base model was approximately 0.63 (95% CI: 0.62–0.66), while for the base model was 0.60 (95% CI:0.58-0.61) (Figure 2B).

**Figure 2:**
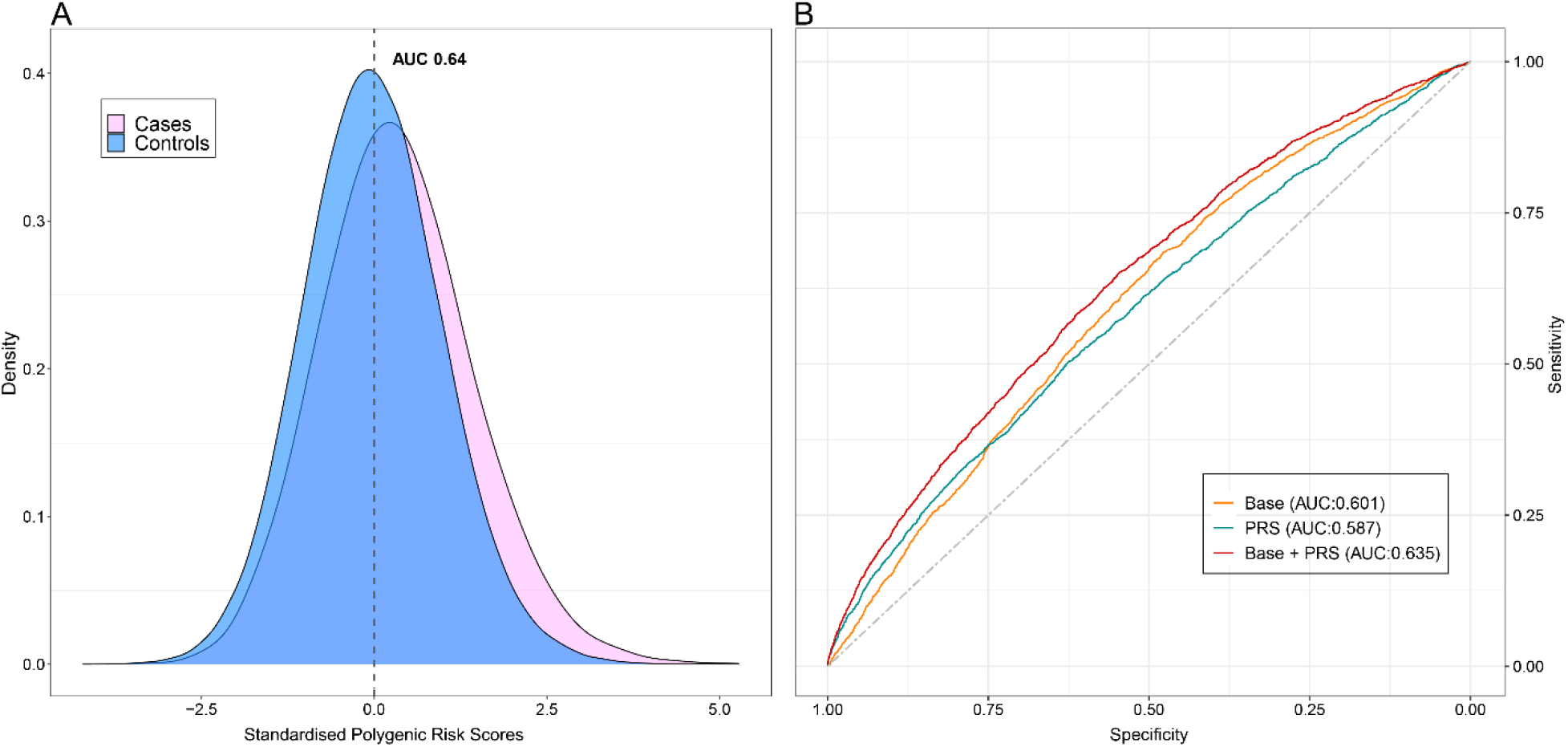
Discriminatory ability of polygenic risk scores in the individuals of the UKB target cohort. (A) Density distribution plot of DVT cases versus controls. (B) ROC curves assess the discriminative power of different models. The grey dot line with an AUC of 50% is used as reference. AUC of the upper red line, showing the combined of polygenic risk category, age and sex is almost 64%. The orange line with an AUC of 60% is calculated based on age and sex factors model, whereas the AUC of the blue line, representing the only contribution from the polygenic risk category, is almost 59%.

We could also observe a clear shift in the distribution of PRSs between the DVT cases and controls of the target cohort (Figure 3, Supplementary Figure 1). Indeed, each standard deviation increase in the PRS was associated with an increased odd of DVT (Table 3). In the target cohort, we could see a clear trend of higher odds of having a DVT diagnosis among individuals of the higher deciles of the PRS (Figure 3): each standard deviation increase in the PRS was associated with an increased odd of disease, reached up 3.12-fold for the highest PRS decile (95% CI: 2.7–3.6; *p* = 5.8×10^−57^). It was clear that the rates of FVL carriers were enriched in the tenth decile (Supplementary Figure 2). To evaluate if the PRS were mainly driven by the FVL and PTM, we also compared the odds of DVT between the PRS deciles when excluding FVL and PTM carriers. Here, there were still 2.3-fold increased odds in the tenth decile (95% CI: 2.03–2.74; *p* value = 9.5×10^−29^) (Figure 3, Supplementary Figure 1).

**Table 3:** The odds ratio among polygenic risk score DVT decile in the UKB. Based on polygenic risk scores, participants were allocated to deciles, from the first decile, the lowest, to the tenth decile, the highest. The first decile was used as reference to the others. The odds ratio and 95% confidence interval were estimated using logistic regression.

| PRS Decile | Number DVT cases | Number controls | Odds ratio | 95% Confidence interval | Odds ratio p-value |
| --- | --- | --- | --- | --- | --- |
| 1 | 270 | 14447 | 1 | 1 | 0 |
| 2 | 310 | 14406 | 1.16 | 0.98-1.37 | 0.08 |
| 3 | 345 | 14371 | 1.30 | 1.10-1.52 | 0.002 |
| 4 | 353 | 14364 | 1.33 | 1.13-1.56 | 0.00047 |
| 5 | 372 | 14344 | 1.40 | 1.20-1.64 | 2.79E-05 |
| 6 | 421 | 14295 | 1.59 | 1.36-1.85 | 4.79E-09 |
| 7 | 446 | 14271 | 1.70 | 1.46-1.98 | 1.26E-11 |
| 8 | 467 | 14249 | 1.76 | 1.52-2.05 | 2.46E-13 |
| 9 | 562 | 14154 | 2.15 | 1.86-2.49 | 2.29E-24 |
| 10 | 796 | 13921 | 3.12 | 2.71-3.59 | 5.79E-57 |

**Figure 3:**
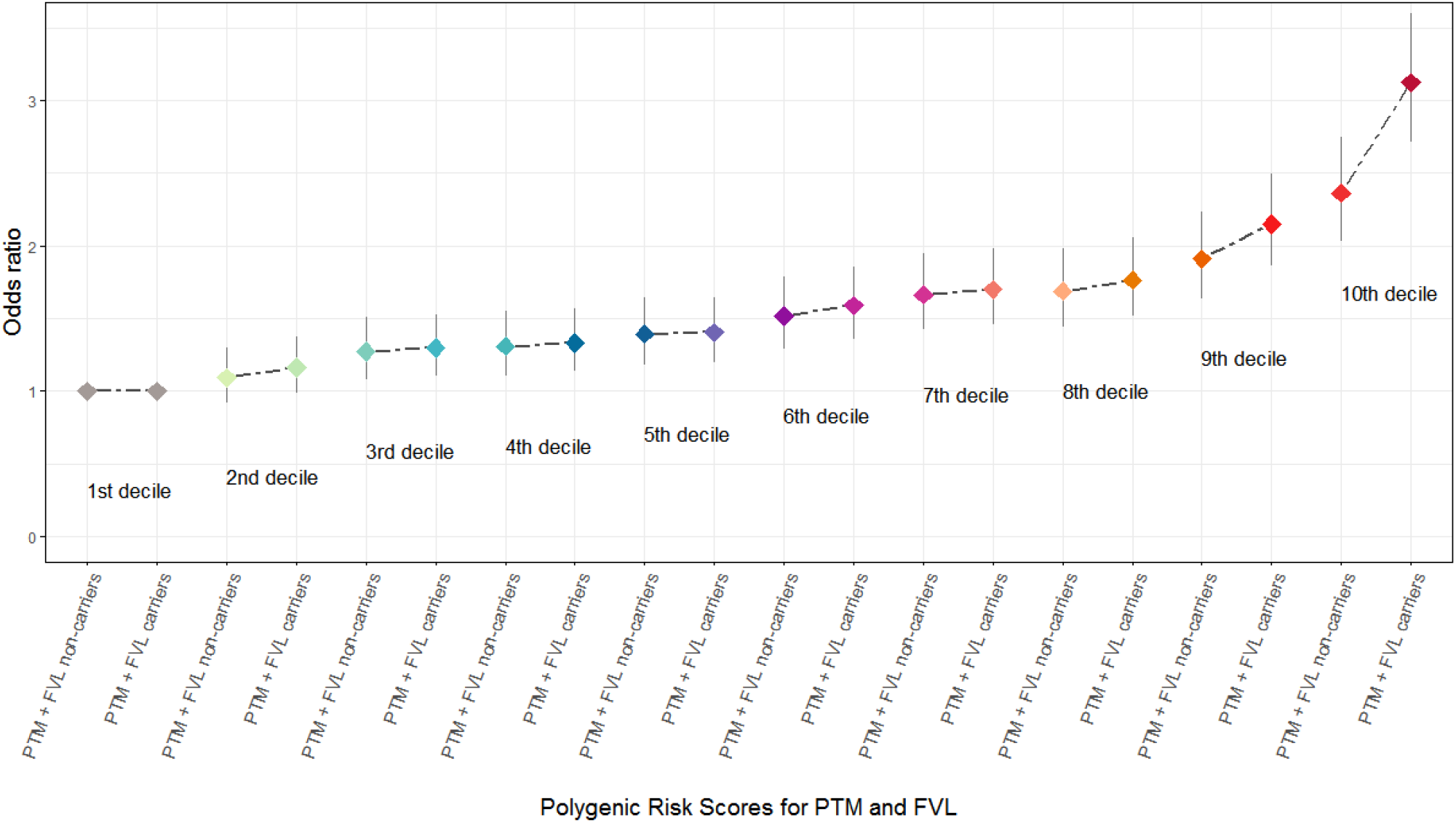
Odds ratio estimates for each polygenic risk score decile with and without carriers of FVL and PTM mutations. Based on polygenic risk scores, participants were allocated to deciles, from the first decile, the lowest, to the tenth decile, the highest. The first decile was used as reference to the others. The odds ratio and 95% confidence interval were estimated using logistic regression. Each point indicates the odds ratios and the bar is lower and upper 95% confidence interval for each odds ratio.

Furthermore, stratifying the cohort by decile of polygenic predisposition we observed a large difference in prevalence of mutation carriers. We found that among DVT cases, the prevalence of FVL and PTM was consistently higher among participants with a high genetic liability of DVT (10% of the cohort), compared with participants having a low genetic liability (10% of the cohort). Specifically, for FVL carriers, the difference in prevalence was 7.6% in the highest decile vs. 0% in the lowest decile. We found that the prevalence of the heterozygotes rare variant in the *CREB3L1* gene (rs187725533) was higher among those with a high polygenic predisposition than those with a low polygenic predisposition (*p* < 0.0008, Fisher’s exact test). Specifically, the difference in prevalence for DVT cases was 2 versus 20, in the lowest and highest polygenic category, respectively (Supplementary Figure 2). Heterozygous carriers of this rare variant in the highest polygenic predisposition were at 2.2-fold (95% CI: 1.34–3.44; *p* = 0.001) increased risk for DVT compared to the lowest category.

### Cumulative incidence

The analysis was performed in 3,735 DVT cases and 141,317 controls. We stratified for the first and tenth decile of the PRS, resulting in in a total of 28,952 participants, of which 936 had a DVT diagnosed. Those in the last decile of the PRS had the highest cumulative DVT risk, of 5% (HR=3.34; 95%CI =2.87–3.88) at 70 years old (Figure 4, Table 4). When zooming on the last PRS decile of risk for those participants carriers of FVL mutations, the predicted cumulative rate risk of DVT was 10%, contraposed to 5% for FVL non-carriers (Figure 5, Table 5).

**Figure 4:**
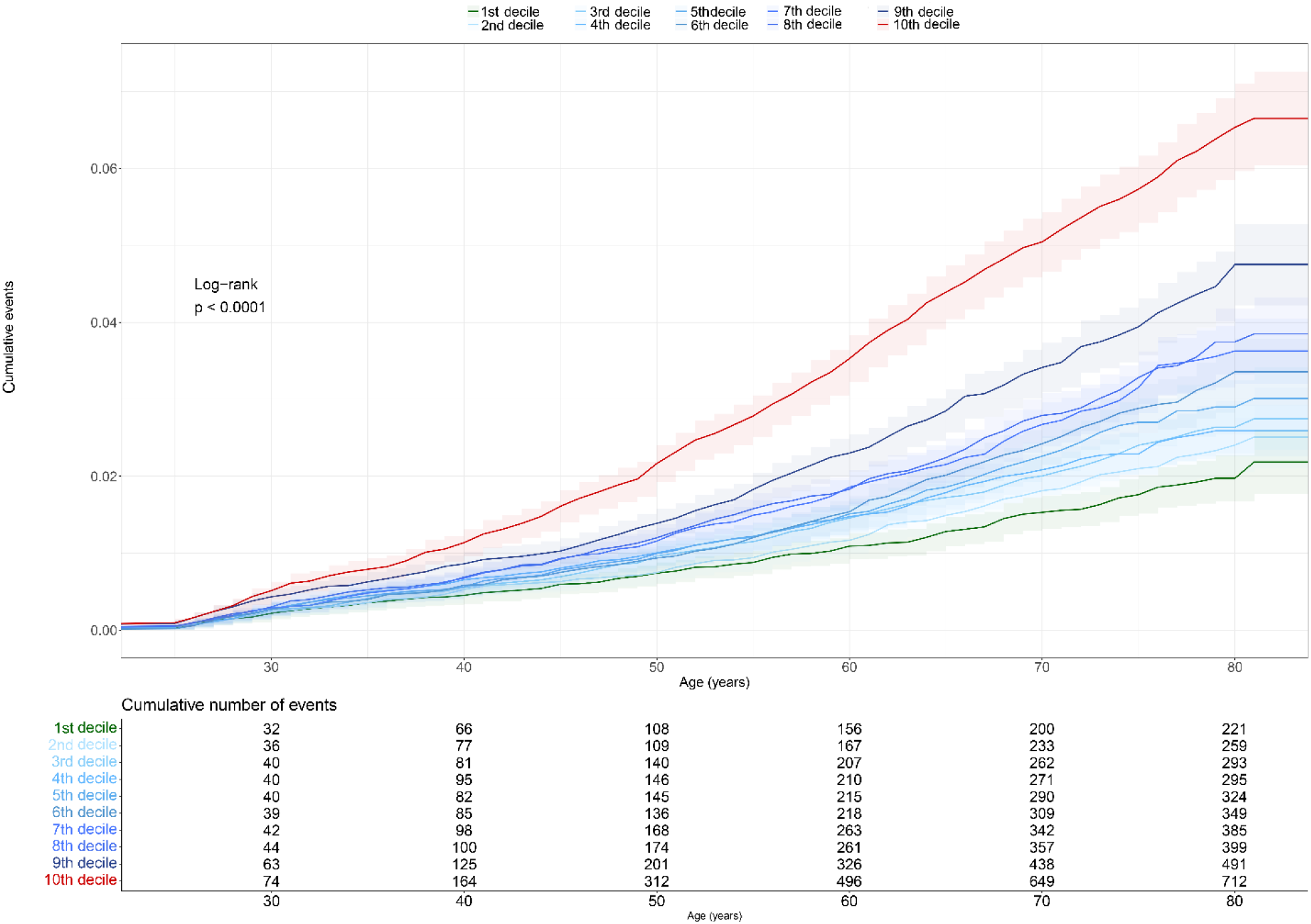
Cumulative event of deep venous thrombosis for each polygenic risk score decile category.

**Table 4:** The hazard ratio among polygenic risk score DVT decile in the UKB. The first decile was used as reference to the others.

| PRS decile | Hazard ratio | 95% Confidence interval | P-value |
| --- | --- | --- | --- |
| 1 | 1 | 1 | 1 |
| 2 | 1.17614 | 0.98-1.40 | 0.075484 |
| 3 | 1.33539 | 1.12-1.58 | 0.001126 |
| 4 | 1.34592 | 1.13-1.6 | 0.000814 |
| 5 | 1.47795 | 1.24-1.75 | 7.04E-06 |
| 6 | 1.58733 | 1.34-1.87 | 7.06E-08 |
| 7 | 1.76206 | 1.49-2.07 | 1.68E-11 |
| 8 | 1.82481 | 1.54-2.15 | 6.15E-13 |
| 9 | 2.255 | 1.92-2.64 | 7.53E-24 |
| 10 | 3.3402 | 2.87-3.88 | 1.13E-55 |

**Figure 5:**
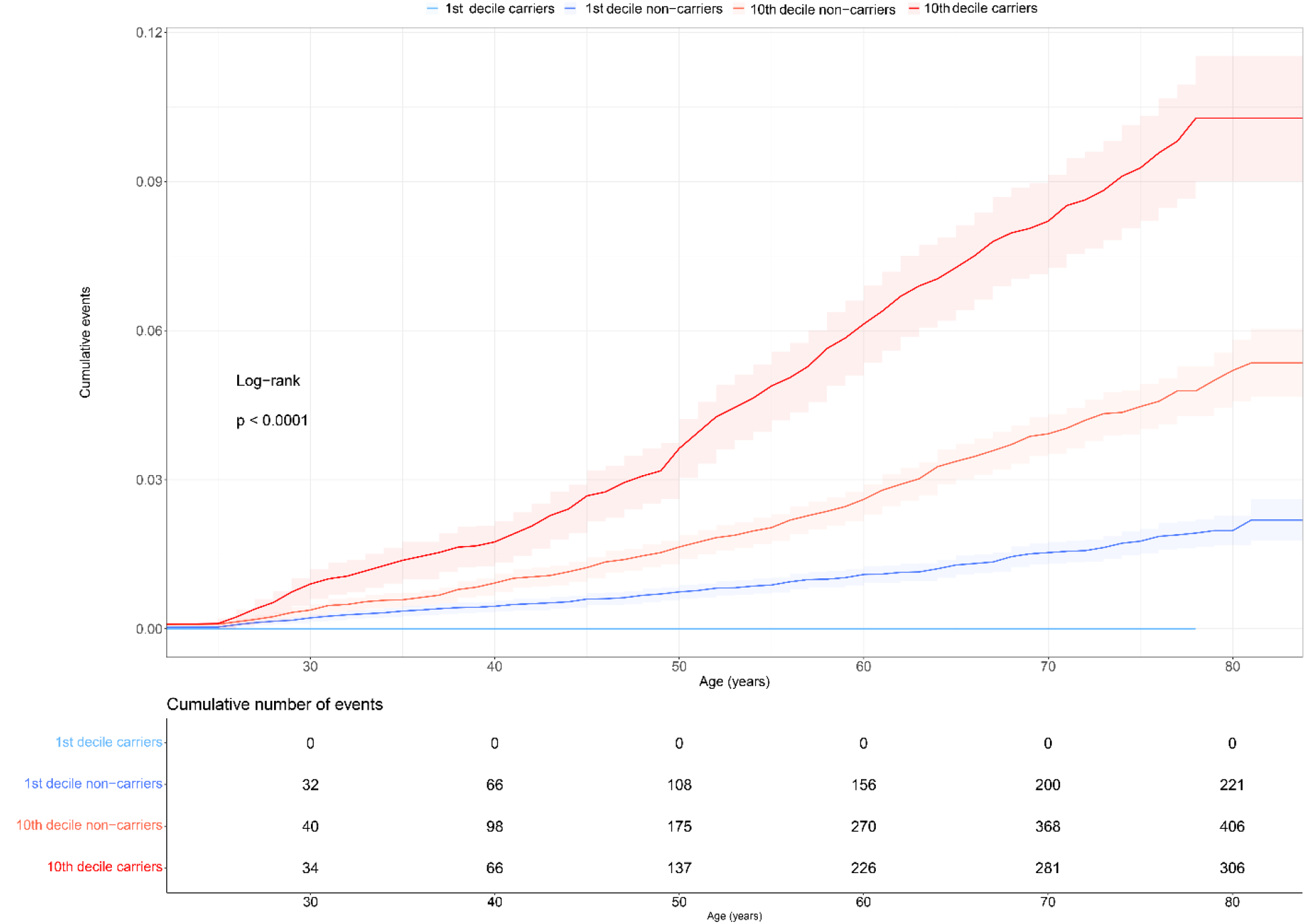
Cumulative event for deep venous thrombosis in comparison between first and last polygenic risk decile categories for FVL mutation carriers and not carriers. The 95% confidence intervals were derived from the survfit object with confidence type set to “log”.

**Table 5:** The hazard ratio between first and tenth PRS decile of risk for those participants carriers and not carriers of FVL. The first decile of not carrier was used as reference to the others.

| PRS decile | Hazard ratio | 95% Confidence interval | P-value |
| --- | --- | --- | --- |
| 1: non-carrier | 1 | 1 | 1 |
| 10: non-carrier | 2.56263261 | 2.17-3.01 | 1.40E-29 |
| 10: carrier | 5.563380006 | 4.68-6.61 | 1.36E-84 |

## DISCUSSION

In this study, we have characterised the genetic contribution of common SNPs and rare variants to DVT, and evaluated DVT incidence rate in participants stratified based on their genetic makeup. We identified and replicated one new common DVT variant (rs11604583) near *TRIM51* (Tripartite Motif-Containing 51) gene, and one novel rare variant (rs187725533) in the *CREB3L1*, the latter associated with as much as 2.2-fold higher risk of DVT.

*TRIM51* is a member of the tripartite interaction motif family of innate immunity regulators and it is predicted to have ubiquitin protein ligase and act as a Ub-ligating (E3) enzyme that takes part in the ubiquitination of proteins.^39,40^ E3 enzymes possess a potential function in the regulation of platelet activation and signal transduction pathways, and platelets constitutively express E3 enzymes.^41,42^ The ubiquitination can be reversed by the action of deubiquitinating enzymes, for example through the editing of the ubiquitin chains. Deubiquitinating enzymes have been detected in platelets. Treatment of platelets with deubiquitinase inhibitors results in diminished thrombin-stimulated platelet adhesion and reduced thrombin-induced platelet aggregation.^43^ Platelet activation has also been suggested to be regulated by the ubiquitin-proteasome system, partially via NF-κB-activation, a key regulator of inflammation.^44^ This indicates that the new DVT locus, *TRIM51*, might be linked to platelet regulation and inflammation through proteasome and E3 ligase activity in the pathophysiology of DVT.

In the exome-sequenced data, the most strongly associated rare variant was the missense variant in *CREB3L1* (cAMP-responsive element-binding protein 3-like 1) gene. *CREB3L1* encodes a transcriptional promoter of arginine vasopressin hormone whose expression is regulated by the cyclic adenosine monophosphate (cAMP) molecule.^45^ Under the positive regulation of cAMP, *CREB3L1* activates the arginine vasopressin promoter that increases the arginine vasopressin hormone expression. *CREB3L1* has been mostly found to be involved in tumour progression in the nervous system and as well to promote vascular smooth muscle cell-neuron interaction.^46,47^ In other studies, *CREB3L1* has also been found to play an important role in maintaining the function of vascular integrity and regulation of the blood pressure.^48^ However, variations in the expression of *CREB3L1* in DVT patients remain to be investigated.

It is possible to reduce mortality by prevention, identifying patients at risk, and understanding the pathophysiology of the disease. Patients with venous thrombosis commonly have an underlying genetic predisposition to thrombophilic disorders that are genetic disorders easily evaluated in a clinical laboratory by DNA analysis or measurement of coagulation factor activity levels. Testing for genetic or acquired thrombophilic disorders has become a common practice in haematology and general medicine, on the basis of the associations of these disorders with risk of a first venous thrombosis. Despite the accumulated evidence supporting the major role played by genetic factors in the pathophysiology of venous thrombosis, only 25% of patients undergoing genetic testing for thrombophilia are carriers of FVL and PTM.^49^ Additional efforts to identify genetic variants associated with venous thrombosis risk are still needed. We therefore developed a PRS to identify individuals with a high genetic liability. Our risk model, in the target cohort and combining the PRS, age and sex, provided adequate power to discriminate participants that developed DVT, with an AUC of approximately 64%. However, it is important to note that interactions between genes and environmental factors can also explain some proportion of individual susceptibility in the development of complex diseases, as shown in the study conducted in young-onset breast cancers women, where it was observed that on the risk of breast cancer, the PRS may interact with hormonal birth control use.^50,51^ Moreover, numerous signals in the loci are within non-coding regions of the genome and due to broad linkage disequilibrium it is difficult to recognize the real genetic cause.^52^

We also observed that the DVT rate was constantly higher in the participants within the highest decile of the PRS across the life-span, and DVT patients with a high polygenic predisposition had a higher likelihood of being heterozygotes of FVL and PTM. In addition, the cumulative risk of DVT at the age of 80 years for FVL carriers in the top PRS decile was 10%, contraposed to 5% for non-carriers. Interestingly, the top PRS decile alone was associated with 3.12-fold risk of DVT, an effect that was still as high as 2.3-fold, when excluding FVL carriers. This clearly demonstrates the need for considering polygenic effects for DVT, and not only well-established mutations such as FVL. These findings imply that PRS may assist in prioritising patients who should triage and undergo sequencing-based genetic tests for the known disease-causing genes. Since current costs for generating a PRS are substantially lower than sequencing, PRS can help to improve the yield of clinical sequencing studies. It is also worth noticing that we did not find any loss of function (LoF) variants in the WES data, in any of the well-known DVT genes, and that both FVL and PTM are common in the population and not classified as LoF. This suggests that the major contribution to DVT might be through polygenic effects of which FVL is contributing to a large degree.

As a result of the ability to identify individuals who are at much higher genetic risk for specific diseases, clinical medicine faces a number of opportunities and challenges. When effective prevention measures are available, attention and resources must be concentrated on those individuals at high risk, integrating genetic risk stratification with other risk variables, such as environmental factors. Where such measures do not exist or are ineffective, identifying high-risk individuals should make it easier to construct effective strategies to find early symptoms. However, risk communication necessitates careful thought. While PRS can be determined at birth, the use of the information and the possible risks to individuals may vary depending on the stage of life.

Our study has several limitations. The PRS described here was derived and tested in participants of white European ancestry from the United Kingdom. Because allele frequencies, linkage disequilibrium, and effect sizes of common polymorphisms vary among ancestries, our PRS will not have optimal predictive power for other ancestries.^53^ Therefore, it is necessary to expand prediction estimates also to non-European ethnic groups. Consequently, additional studies are warranted to validate risk estimates within other populations. We also constructed the DVT risk prediction model including PRS, age and sex, obtaining an AUC of almost 0.64. Our discriminatory accuracy is lowered compared with the combined risk assessment model (genetic and clinical factors, such as cancer, recent hospitalization, use of oral contraceptive pills, and obesity) reported by Kolin^54^ (AUC of 0.7) in patients affected by VTE, defined as deep vein thrombosis and/or pulmonary embolism, but higher of the score calculated by De Haan (AUC of 0.54).^55^ Last, we did not account for gene and environment interactions, and as a result, SNPs that interact with environmental factors may be undetected if they don’t have main effects or if their interaction effect is in a different direction. We should also highlight that several of the participants did not have a date of the first DVT diagnosis available and were therefore excluded from some of the analyses. Consequently, the DVT rates estimated in this study are lower than in the population in general. However, there is no reason to believe that this bias is correlated with the genetic liability to DVT, and the rate ratios and difference between PRS deciles are therefore likely to be generalizable to the UK population.

In conclusion, we showed that common and rare variants influence DVT risk, and that the PRS improves risk prediction on top of FVL and PTM. This suggests that individuals classified with high PRS scores could benefit from early genetic screening, and that the diagnostics should take advantage of the genetic screening for well-known mutations to evaluate the individual risk. Further evaluations of the genetic risk variants in other ancestries are warranted.

## Data Availability

All data produced in the present study are available upon reasonable request to the authors.

## Acknowledgements

We acknowledge all of the participants and staff involved in UKB for their valuable contribution. The computations were performed on resources provided by Swedish National Infrastructure of Computing (SNIC) through Uppsala Multidisciplinary Center for Advanced Computational Science (UPPMAX) under projects SNIC 2018/8-372 and sens2017538.

