## Supplementary Figure for "Using genotyping and whole-exome sequencing data to improve genetic risk prediction in deep venous thrombosis"

### Supplementary figures

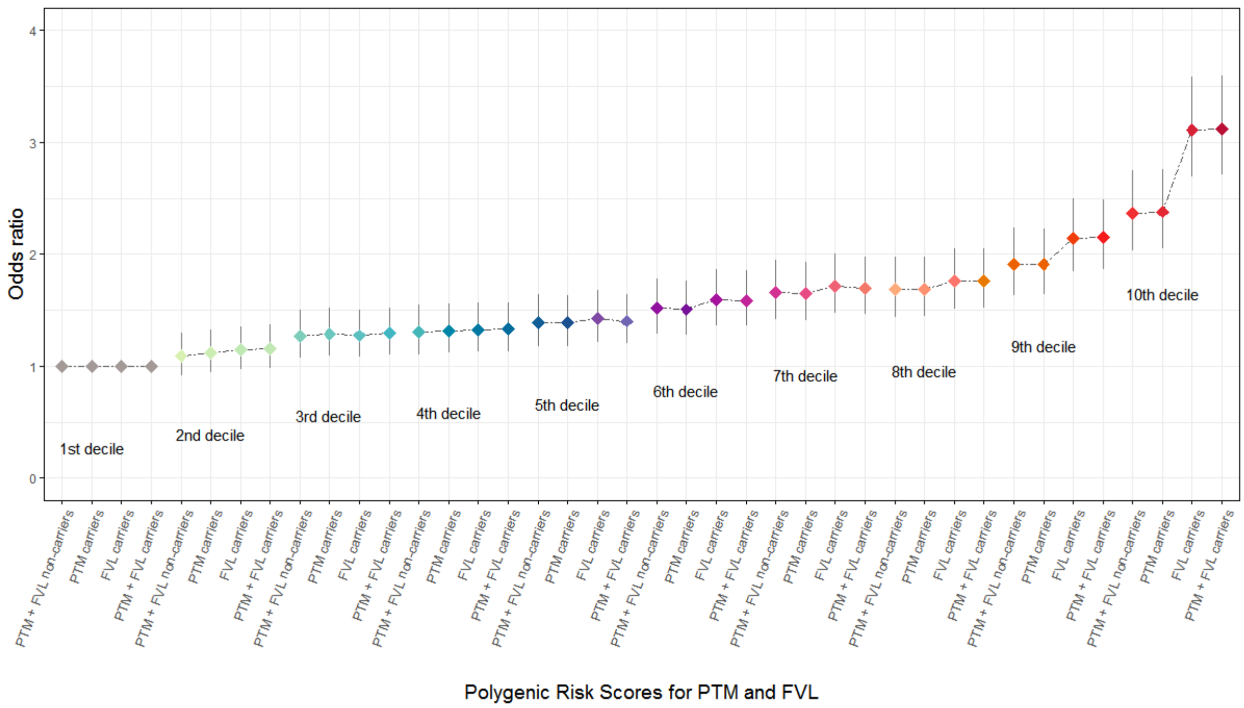


**Supplementary Figure 1: The odds ratio for carriers of risk alleles of FVL and PTM mutations, or both, for each polygenic risk category.** The first decile was used as reference to the others. Each point indicates the odds ratios and the bar is lower and upper 95% confidence interval for each odds ratio.

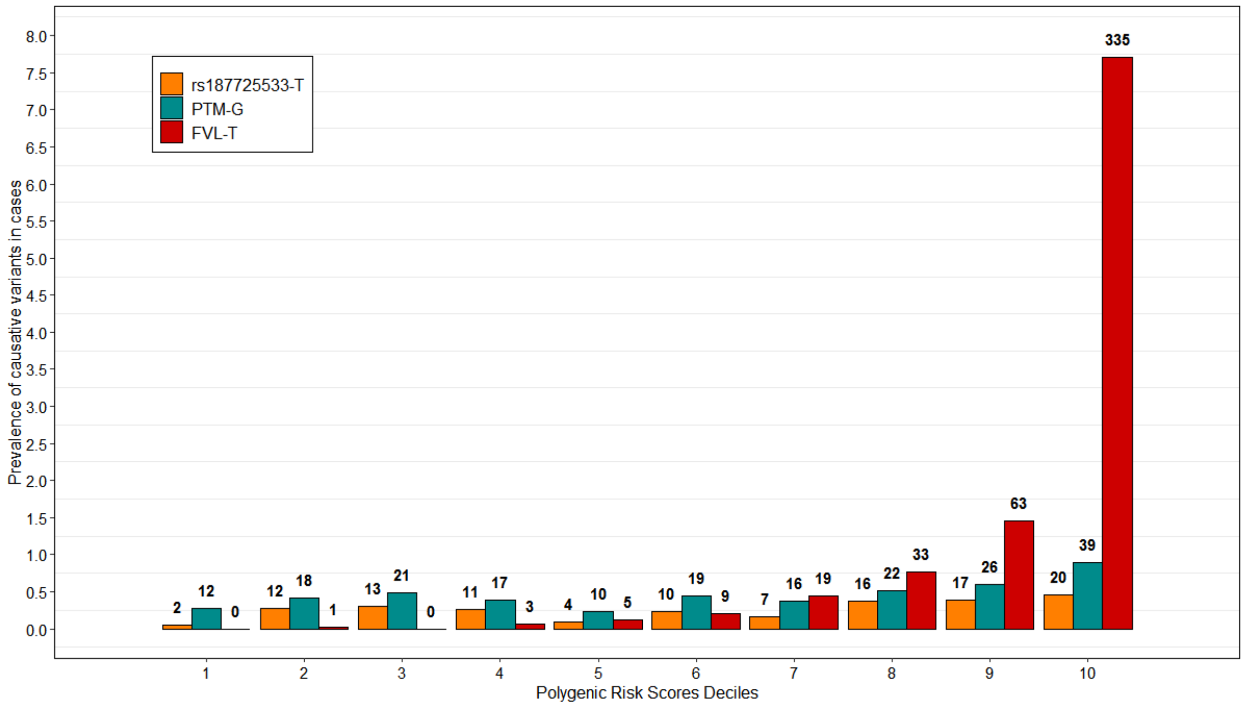


**Supplementary Figure 2: Absolute frequencies of risk alleles for rs187725533 variant in *CREB3L1,* PTM and FVL mutations in deep venous thrombosis participants for each polygenic risk category.**
